# Experimental study on killing ticks in wild natural environment

**DOI:** 10.64898/2026.03.09.26347948

**Authors:** Yan-Dong Wang, Shun-Shuai Liu, Yi-Chuan Yang, Jun Du

**Author notes:** **Address for correspondence author**: Ju Du, Zibo Center for Disease Control and Prevention, Zibo, China;.

## Abstract

A field trial was conducted using 10% lambda-cyhalothrin microcapsule suspension to provide a method for killing ticks and preventing diseases in outdoor gatherings of people or temporary resettlement places after disasters. In this study, three field experimental sites were selected, and each experimental site was set up with a test area and a control area. Before pesticide application, the tick density in three test areas and three control areas was surveyed using the flagging method. Subsequently, two methods were used for pesticide spraying: motorized fogging and electric constant-volume spraying (with the pesticide diluted 300 times). The relative density decline rate of ticks was calculated in three test sites on days 1, 7, 14, 21, and 28 after spraying, and all experimental areas achieved good tick-killing effects. Even without prohibiting wild animals, grazing sheep, and dogs (which are often infested with ticks and not treated) from entering the trial sites, spraying 10% lambda-cyhalothrin microcapsule suspension could maintain a tick-free (low-density) state for approximately 3-4 weeks. Our study provides an idea for controlling epidemics through tick elimination during the high incidence period of tick-borne diseases.

## Introduction

Ticks are blood-sucking arthropods that are widely distributed around the world. From 2005 to 2020, the number of reported cases of vector-borne infectious diseases in China showed a fluctuating downward trend, while the number of cases of Severe fever with thrombocytopenia syndrome (SFTS) showed a significant upward trend [1], ticks are the main vector of SFTS [2]. Ticks have a wide range of host selectivity and the ability to efficiently spread pathogens, making them an important vector after mosquitoes [3]. Ticks can spread viruses, bacteria, and protozoa [4], posing a serious threat to humans and animals. Ticks live in a wide range of environments, such as forests, grasslands, shrubs, and even urban parks [5], although the current research on ticks, tick-borne diseases and the pathogens they carry has been studied deeply, in the wild, especially in the vast rural areas, the protection awareness of tick bites is relatively weak. Compared with a series of pathogen screening and diagnosis after tick bites, reducing tick density and preventing tick bites is also a disease prevention idea. Zibo City is located in the central region of Shandong Province in the East China, with many mountainous hills in the south of the city (Figure 1), the density of ticks in the wild is high, and it is a high incidence area of tick-borne diseases.

**Fig. 1.**
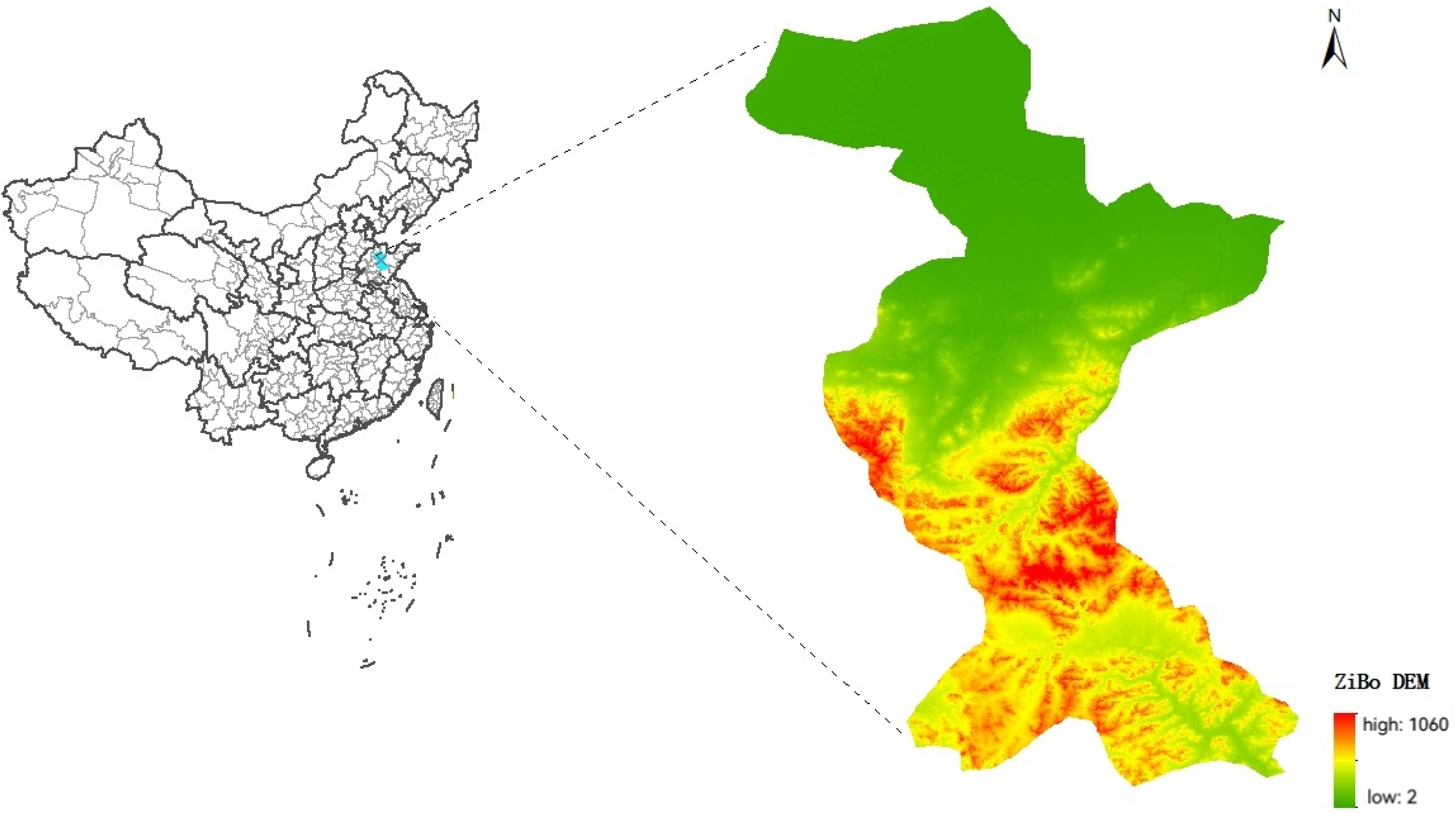
Topography of Zibo City

In this study, the field killing effect of a sanitary insecticide on ticks was reported for the first time. Lambda-cyhalothrin is an efficient, broad-spectrum and quick-acting pyrethroid insecticide, which has the effect of avoiding, knocking down and poisoning insects [6,7]. Microcapsule suspension is an advanced dosage form that encapsulates the active ingredient in the polymer capsule wall and disperses in water. It has the advantages of sustained release, improved stability and reduced toxicity [8]. We demonstrated that 10% lambda-cyhalothrin microcapsule suspension for killing ticks in the field can maintain a tick-free (low density) state for about 3-4 weeks, which is of great significance for controlling the epidemic situation in the high incidence period of tick-borne diseases.

## Materials and Methods

### Test sites

Three vegetation-rich hillside fields in Zichuan County, Zibo City, Shandong Province were selected as test areas for tick control with tickcide. Filed 1 to filed 3 were 700 square meters, 1000 square meters, and 1000 square meters, respectively. Field 3 consisted of two areas with one area of 800 square meters (Area A) and another area of 200 square meters (Area B). The 3 fields were used as test areas for tick control by spraying tickcide, and a control area with approximately the same area as the test area was set up around each test filed. Each test area and its control area were separated by 5 meters to prevent drug dispersion. The three test areas were separated by more than 300 meters. All six test and control sites meets the basic requirements of a baseline tick density of ⩾ 10 ticks (each flag per hour). The test area was sprayed, and the control area remained in a natural state without any treatment. The vegetation, light, temperature and humidity of all six test areas were similar.

### Application equipment and tickicides

The 10% lambda-cyhalothrin microcapsule suspension (Jiangsu Gongcheng Bio-tech Co. Ltd.) was used as a tick-killing reagent, and the electric constant sprayer (Suzhou Taiyuwei mechanical and electrical technology Co. Ltd.) and the motorized mist sprayer (ANDREAS STIHL AG&CO.KG) were used for spraying.

### Application method and drug dosage

The experiment began in mid-July 2025. The weather was sunny on the day of application. The electric constant sprayer was used in No.1 field, the motor mist sprayer was used in No.2 field, the motor mist sprayer was used in A area of No.3 field, and the electric constant sprayer was used in B area. The average application amount was 60 ml/m^2^ and the drug concentration was 0.03 % (10% lambda-cyhalothrin microcapsule suspension diluted 300 times). The method and time of application are detailed in table 1.

**Table 1.** Field spray of tickcide.

| Experimental sites | Area(m<sup>2</sup>) | Application time (h) | Application dose (%) | Application dosage (L) | Application Equipment |
| --- | --- | --- | --- | --- | --- |
| No.1 site | 700 | 15: 25~16: 00 | 0.03 | 42 | Electric constant sprayer |
| No.2 site | 1000 | 10: 00~10: 40 | 0.03 | 60 | Motorized mist sprayer |
| No.3 site (A) | 800 | 11: 20~12: 00 | 0.03 | 48 | Motorized mist sprayer |
| No.3 site (B) | 200 |  | 0.03 | 12 | Electric constant sprayer |

### Tick monitoring

The density of questing ticks was monitored before and after the tickcide spray. The ticks were dragged for 0.5 h in the test area using a cloth flag method, and the collected ticks were loaded into the test tubes for classification and identification, and the density calculation. Tick density was calculated with a formula of D = N / T; D is the density of ticks, the unit is numbers of ticks/each flag per hour); N is the total number of ticks collected; T is the time of collecting ticks, in hours (h).

### Verification of tick killing effect

The density of questing ticks in the test area and the control area of each field was monitored by flagging on day 1, 7, 14, 21, and 28 after spraying, respectively. The changes of tick density before and after spraying were compared, and the relative density decline rate of ticks was calculated. The relative density decline rate (%) = (A-B) / A × 100 %, A is the density of ticks in the control area, and B is the density of ticks in the spraying area. The effect was significant when the relative density of ticks in the application area decreased by ≧ 80 % on the first day after application, and the monitoring of the follow-up time was used to observe the duration of the efficacy.

## Results

### Background density of questing ticks in the test sites

The average density of ticks in No.1 field area was 238 / (each flag per hour), and that in control area was 95 / (each flag per hour). The average density of ticks in the No.2 field area was 28 / (each flag per hour), and that in the control area was 24 / (each flag per hour). The average density of ticks in the No.3 field area was 271 / (each flag per hour), and that in the control area was 76 / (each flag per hour).

### Tick density monitoring after pesticide application in the experimental area

The density of questing ticks was monitored on day 1, 7, 14, 21 and 28 after spraying of pesticie in the fields. The ticks captured in the test areas and the control areas were classified and identified as *Haemaphysalis longicornis*. The number of ticks in the No.1 test area was 0, 12, 0, 0, 38 / (each flag per hour), and in the control area was 42, 108, 140, 72, 56 / (each flag per hour). No ticks were captured in the No.2 test area, and the control area was 24, 34, 54, 4 and 18 ticks / (each flag per hour). No ticks were captured in the No.3 test area A. The number of ticks in No.3 area B was 0, 0, 0, 2, 1 / (each flag per hour), and the control area was captured 117, 116, 119, 71, 65 / (each flag per hour), as shown in Fig.2.

**Fig. 2.**
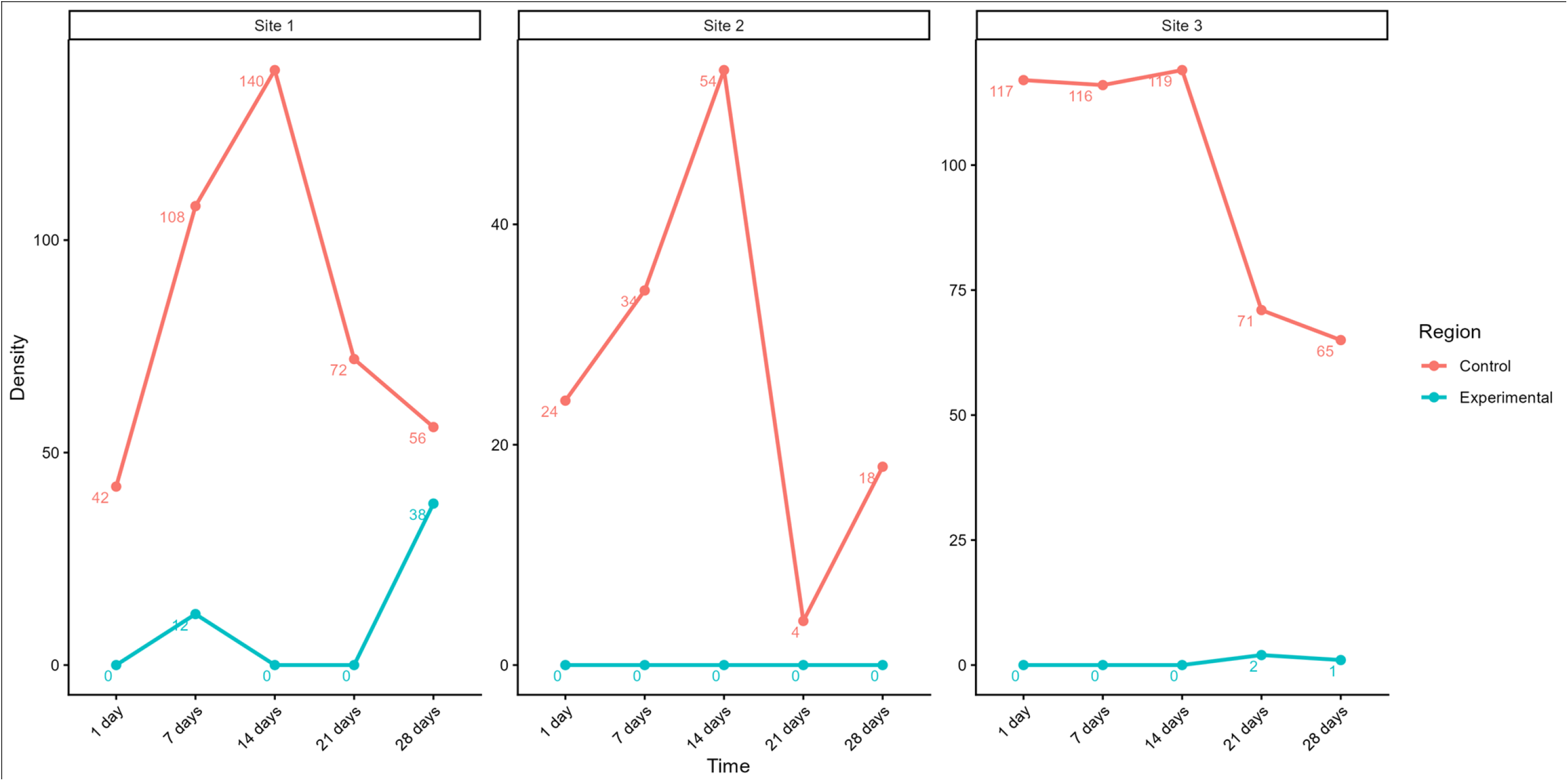
Monitoring of tick density in the testing areas

The killing rate of ticks was 100 % on the first day after application. After 7, 14, 21 and 28 days, the density of ticks in the application area was smaller than that in the control area. Until the 28th day, the relative density of ticks in No.1 was decreased by 32.1%, which was less than the set target value of 80%.

## Discussions

The 10% lambda-cyhalothrin microcapsule suspension was used for the first time in the field natural environment tick control test, and it had a good control effect on *Haemaphysalis longicornis* (the killing rate was 100% on the first day). After continuous observation for one month, the tick density in the three test areas remained at a low level, indicating that the use of the pesticide for a tick killing operation can suppress the tick density in the area for 3 to 4 weeks. It has reference value for tick killing and disease prevention in field population gathering activities and post-disaster field temporary resettlement places, and also provides basic data for expanding the application scenarios of these sanitary insecticides. This is consistent with the test results of the two insecticides on the killing effect of ticks in 2024 [9]. This result provides a new idea for the elimination of ticks in the main activity sites during the high incidence of tick-borne diseases, reducing the risk of tick bites on humans, and thus reducing the occurrence of diseases.

At present, there are few reports on the research of tick killing agents in the field, such as cis-cypermethrin suspension [9], and the Credelio Quattro carried out in the laboratory to kill *Haemaphysalis longicornis* [10], but few reports on the actual effect of killing ticks in the field environment. There is a pesticide quasi-brand tick killing agent, which is used to kill ticks on the surface of mammals such as sheep and cattle. This report showed that the effect of 10 % lambda-cyhalothrin microcapsule suspension showed sound effectiveness and persistence, and the advantages were significant. First, efficient, the action time is short, the density is rapidly reduced, and only one application is required; Second, the effect is lasting, and the tick killing effect can be maintained for 3 to 4 weeks; Third, environmental protection and low toxicity, using microcapsule technology to reduce environmental pollution, safety and environmental protection.

The limitations of this study include: First, with the extension of time, the ticks in the control area will decrease with the increase of the number of ticks, and the tick density in the control area will be artificially reduced. There is a deviation in the statistics of the results. If the density of ticks selected in the test plot is low, it may affect the judgment of the results. Second, the test area is an open environment, and there will be sheep passing. Wild animals such as rodents and birds also exist. The introduction of new ticks cannot be avoided, and the test results may have an impact. Third, the test time was selected at the peak of tick density, which was also in summer. The precipitation was large, and a large amount of water in the mountains may have an impact on the density of ticks. Before the density was monitored on the 21st day, it had just rained heavily. There were signs of sediment erosion in the test area and the control area (No.1 and No.2 plots were more obvious). The density of ticks in the control area was also relatively low.

The type of sprayer may affect the efficacy (uniformity, penetration); the results showed that ticks were detected on the 7th, 21st and 28th day after spraying with constant sprayer in No.1 test area and No.3 test area B, respectively. The area where ticks were detected on the 7th day in No.1 test area was at the edge of the plot, with more weeds and low shrubs. The relative density of No.1 test area decreased by 32.1% on the 28th day, and the effect was not obvious. The penetration of the constant sprayer is poor, and the droplets can only reach the upper surface of the leaves. The droplet of the motor mist machine is smaller, the air volume is larger, and the weed leaf surface will be blown and oscillated. When spraying, it can cover the leaf surface, leaf bottom, and ground [11], which can fully kill the ticks in this area and maintain the efficacy for a longer time. In addition, it takes 12-14 min for the electric constant sprayer to apply the same amount of 12L, which can cover the area of 2-3 m, while it takes 4-5 min for the motorized mist sprayer to cover the area of 4-5 m, which saves more time and labor.

In summary, the prevention of tick-borne infectious diseases, the reduction of tick density, and the prevention of tick bites are the key. In the high-density season of ticks, targeted tick control interventions are carried out in local areas to rapidly reduce the density of ticks and maintain a low density for a long time, reducing bites and declining the probability of disease occurrence. This idea requires more discussion and practice.

## Data Availability

We declare that our manuscript involved complete data and no additional data are available

## Acknowledgment

the authors are grateful for Professor Xue-Jie Yu, Wuhan University for discussion of the manuscript.

## Author contributions

Experimental design, conceptualization, data summary statistics, manuscript correcting: Yan-Dong Wang, Jun Du. Writing-original draft, data analysis and visualization: Shun-Shuai Liu. Experiment: Yi-Chuan Yang. Writing-review and editing: Xue-Jie Yu. All authors read and approved the manuscript.

## Funding

There is no funding.

## Availability of data and materials

We declare that our manuscript involved complete data and no additional data are available.

## Declarations

### Ethics approval and consent to participate

No applicable.

### Consent for publication

All the authors consent to the publication of the manuscript.

### Competing interests

The authors declare no competing interests.

## Notes

### Competing Interest Statement

The authors have declared no competing interest.

### Funding Statement

The author(s) received no specific funding for this work.

